# Prospective Evaluation of a 90-day Mortality Prediction Model: From Silent Pilots to Real Time Deployment in the EHR^*^

**DOI:** 10.1101/2023.01.25.23284977

**Authors:** Lorenzo A. Rossi, Laura Roberts, Finly Zachariah

**Affiliations:** City of Hope National Medical Center, Duarte, CA, USA

## Abstract

Prognostication in oncology is increasingly difficult due to the rapid evolution of therapies with significant improvement of survival. Accurate prognostication is essential to provide optimal, value-driven end of life care for cancer patients, and can promote goals of care (GOC) conversations with the potential to minimize chemotherapy or ICU utilization in the last weeks of life, and possibly increase hospice admission and length of stay.^1^ There are several recent publications on the application of machine learning for prognostication.^2,3^

We developed a 90-day mortality prediction model trained with data in the Electronic Health Records (EHR). After a non-interventional pilot stage, we deployed the model in February 2021 in the real-time Electronic Health Record Epic infrastructure of our cancer center. Here we present the model and evaluate its overall performance for the first 7.5 months since the go-live and outline our evaluation process for the next stages.

## Methods

We trained a gradient boosted tree classifier (via the XGBoost library) with observations from 28, 484 patients (less than 10% non-oncologic) and 493 features from demographics, lab test results, flowsheets and diagnoses collected from the EHR in our medical center between 2014 and early 2019. We extracted several hand-crafted features from the time series of labs and flowsheet data in the 180-day temporal window preceding the time of each prediction. We imputed missing values for features only in obvious cases, because tree-based classifiers can handle missing data. Table 1 lists the clinical variables used by the model and the type of features extracted from lab and flowsheet time series. The features associated with the diagnoses consisted of aggregations of Word2Vec embeddings of the ICD-9 codes. A portion of the observations were used for retrospective evaluation based on a temporal split to mimic real world deployment, where past observations are used to train a model to predict in the present. For inclusion in training and evaluation sets, patients needed to have at least two encounters and alive patients needed to have an active encounter in the EHR at least one year after the prediction date. To limit risk of leakage, we picked dates of prediction to exclude encounters within 7 days of death. We also avoided over-representing observations with prediction dates within 30 days of death to avoid training the model with a disproportionate number of near-term deceased patients.

**Table 1:**
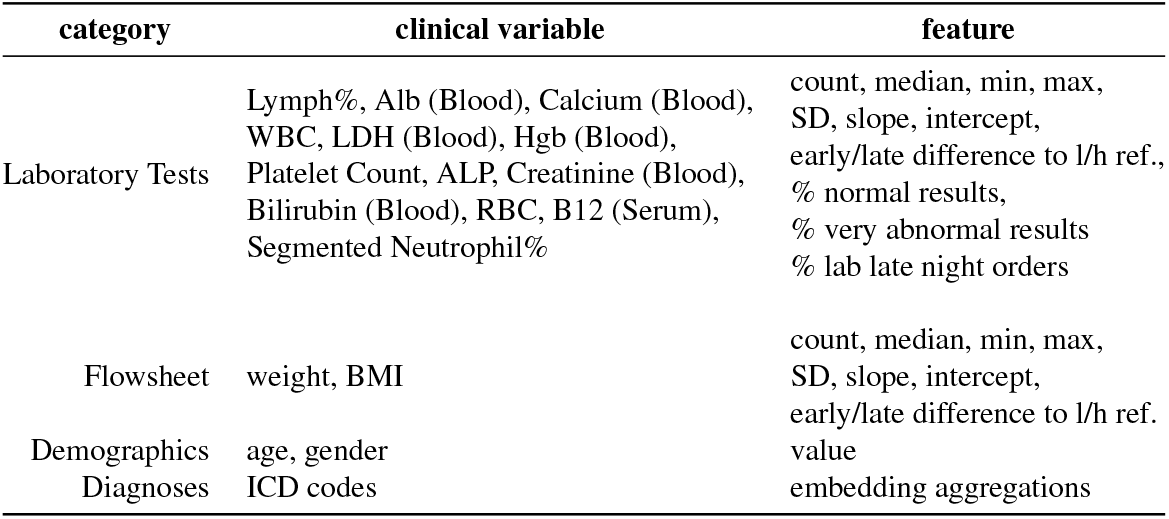
Types of clinical data used to extract the features for the machine learning models.

After hyperparameter tuning via cross-validation and retrospective evaluation, we retrained a version of the 90-day mortality model for deployment in a silent (invisible to clinicians) prospective pilot, inclusive of all City of Hope patients with accessible EHR data. The resulting model consisted of an ensemble of 413 decision trees with 6 maximum depth. Since October 2019, the model made batches of predictions from observations automatically queried once a day from our electronic data warehouse (EDW). The presence of select new lab results triggered a prediction. We ran the pilot for the entire 2020, retraining the model several times and then deployed the model in the medical center’s EHR real time infrastructure (RTI) in February 2021, In the RTI, the model receives a stream of Health Level 7 (HL7) messages. Pilot and production included predictions for both inpatients and outpatients.

## Results

In the retrospective evaluation (*n* = 5, 037), areas under receiver operating characteristic (AUROC) and precision-recall (AUPRC) curves were 0.82 and 0.20 respectively with 5% prevalence. Figure 1 reports ROC and PRC curves, and performance metrics for pilot (year 2020) and real time production implementations (February 15 deployment - September 30, 2021). The prevalence is the 90-day mortality rate. Predictions are triggered by new recordings of certain lab results. Hence, there may be multiple predictions for a patient. The median age of the patient population at the time of first prediction in production was 52 years (18-101 years range). The performance for pilot and production implementations were very consistent: 0.86 (95% CI: 0.860 − 0.864) v.s. 0.85 (95% CI: 0.844 − 0.849) AUROC and 0.41 (95% CI: 0.41 − 0.417) vs. 0.39 (95% CI: 0.39 − 0.402) AUPRC, respectively. The lead days are the time between a correct mortality prediction and the death date of a patient.

**Figure 1:**
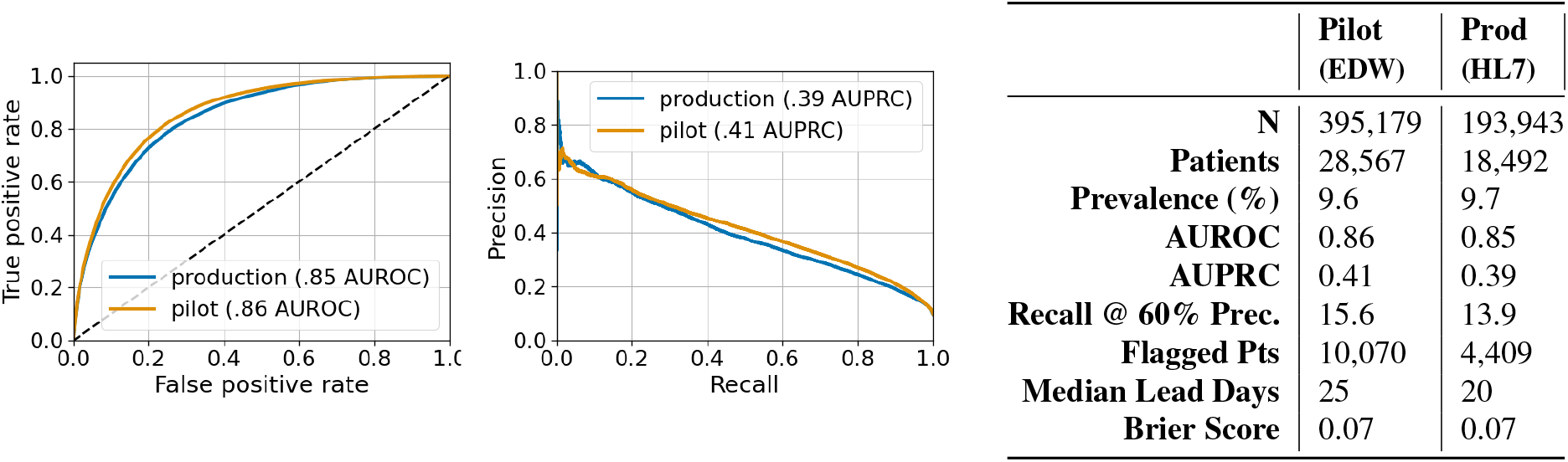
ROC and PRC plots; Table with pilot and production performance metrics.

## Discussion and Outline for Next Stages of AMIA AI Evaluation Showcase

A common pitfall for deployments of machine learning models in production is a drop in performance with respect to previous evaluations. In our production deployment, overall performance was consistent with the 2020 pilot and the retrospective evaluation at development stage. Pilot and production relied on different data infrastructures: EDW vs. real time streams of HL7 messages.

The production model is being employed in an increasing number clinical workflows: e.g. to help Clinical Social Work to prioritize patients for advance directive completion, to identify patients who would benefit from GOC discussions, to order Supportive Care consults for ICU transfers of high risk patients, and to identify patients for trial recruitment.

In Stage II, we will present in detail the different clinical decision support applications of the model with related workflows and specific performance results. We will include also results from clinician surveys on utility and usability.

In Stage III, we will report results on our quality measures (e.g. completion rates of advance directives and GOC conversations), as well as on national end of life quality measures (e.g. time in the ICU in the last 30 days of life, chemotherapy use in the last two weeks of life, and hospice utilization metrics).

## Data Availability

The data associated with the current study is not publicly available as the data are not legally certified as being deidentified. Summary data
may be available by contacting the corresponding author.

## References

1. Scott D Halpern. Goal-concordant care-searching for the holy grail. The New England Journal of Medicine, 381(17), 2019.

2. Kenneth Jung, Sylvia E. K. Sudat, Nicole Kwon, Walter F. Stewart, and Nigam H. Shah. Predicting need for advanced illness or palliative care in a primary care population using electronic health record data. Journal of biomedical informatics, 92, 2019.

3. Vincent J Major and Yindalon Aphinyanaphongs. Development, implementation, and prospective validation of a model to predict 60-day end-of-life in hospitalized adults upon admission at three sites. BMC medical informatics and decision making, 20(1):1–10, 2020.

